# MAM: Flexible Monte-Carlo Agent based Model for Modelling COVID-19 Spread

**DOI:** 10.1101/2022.09.11.22279815

**Authors:** Hilla De-Leon, Dvir Aran

## Abstract

In the two and half years since SARS-CoV-2 was first detected in China, hundreds of millions of people have been infected and millions have died. Along with the immediate need for treatment solutions, the COVID-19 pandemic has reinforced the need for mathematical models that can predict the spread of the pandemic in an ever-changing environment. The susceptible-infectious-removed (SIR) model has been widely used to model COVID-19 transmission, however, with limited success. Here, we present a novel, dynamic Monte-Carlo Agent-based Model (MAM), which is based on the basic principles of statistical physics. Using data from Israel on three major outbreaks, we compare predictions made by SIR and MAM, and show that MAM outperforms SIR in all aspects. Furthermore, MAM is a flexible model and allows to accurately examine the effects of vaccinations in different subgroups, and the effects of the introduction of new variants.

## Introduction

In the 18th century, Swiss mathematician Daniel Bernoulli developed mathematical models to study how variolation could be used to control smallpox [1]. Since then, researchers have used many approaches to develop models that can examine and explore the dynamics of infectious disease transmission. Nowadays, mathematical modelling of disease spread has become essential for decision-making on the national and international level, especially during the COVID-19 pandemic [2-5]. Globalization had, however, changed geography, social conditions, and transportation across countries, even compared to twenty years ago when SARS-CoV-1 was first detected. Consequently, mathematical models need to be adapted to a world that is not static but varies from place to place and over time.

The main metric for assessing the spread of a disease is *R*, the reproduction rate; when *R* is above 1, one individual infects, on average, more than one other individual, which indicates the disease is spreading. A virus usually is represented by the basic reproduction rate, ***R***_**0**_, the transmission rate without any measures, while we denote with ***R***_***e***_ the observed reproduction rate in the population]. ***R***_***e***_ is affected by pharmaceutical and non-pharmaceutical interventions (NPIs), and by the level of protection against infection in the population (i.e., the recovered and vaccinated populations). It is important to note that there are many factors that affect assessment of ***R***_***e***_, and for example, testing policies, which vary across countries over time, directly impacts the assessment of ***R***_***e***_. Thus, relying solely on daily case numbers as a measure of spread is problematic, especially when comparing spreads among different countries.

The availability of vaccines, which started by the end of 2020, add another layer to the complexity of assessing spread. We now differentiate between ***R***_***t***_ the theoretical reproduction rate of the virus, and ***R***_***e***_, the effective reproduction rate of the virus. ***R***_***t***_ estimates the rate of encounters between infected and non-infected individuals that would have resulted in an infection if the vaccines had not been available. ***R***_***e***_, on the other hand, is affected also by vaccination rates and the vaccine’s effectiveness. Since vaccination rates vary not only internationally but also by age group, and since vaccine effectiveness decreases over time, and different variants differ in their resistance to the vaccine, one needs to consider all the above when attempting to predict ***R***_***e***_. Such predictions must consider the unique conditions in the country, including the dominant variant, the current restrictions (e.g., NPIs), the vaccination rate for various age groups, and the effectiveness of the vaccine.

There are two main approaches to model disease spread: empirical and mathematical models. Empirical, or statistical, models use available data and statistical and machine-learning-based approaches to make predictions on future spread dynamics or the temporal evolution of the disease in terms of severe morbidity [6-13]. The main drawback of such models in COVID-19 is their inability to predict future outbreaks in the presence of changing conditions, for example, a scenario of a new variant or the vaccination of the population. Statistical models, however, had a major role in clinical outcome predictions of COVID-19 patients since disease progression was associated with individuals’ health conditions more than with the particular variant [14].

Mathematical models are sets of coupled differential equations to predict the spread of disease. The SIR methodology [15] and its refinements, such as the SEIR model [16] are the main known examples of this class. They have been the dominant approach in the scientific literature for studying infectious diseases and have been applied widely over the last decades. In general, SEIR stands for “Susceptible, Exposed, Infected, Removed,” which serves to decompose the population. The model describes an epidemic via the movement of the population from one compartment to another. Susceptible individuals become exposed, infected, and finally removed from the population. Removal can be either by recovery or by death. Transitions from one state to another are governed by rate equations. According to the SIR model, the spread rate is determined primarily by ***R***_***e***_, the effective reproduction rate, where there is a population of susceptible individuals, and the number of infectious cases directly resulting from one infected individual is ***R***_***e***_.

One disadvantage of population-level mathematical models is that they are limited in the ability to model system dynamics, especially when various population subgroups have different dynamics. We argue that a model that can accurately predict the spread must be able to model subpopulations that may have different spread dynamics. Since Nonpharmaceutical Interventions (NPIs) are essential in controlling COVID-19, a geography-based model is necessary because these NPIs fluctuate across regions and countries. Also, a successful model should divide the population into several age groups, matching their varying patterns of social interactions. A realistic model would require writing a different equation for each subgroup, which complicates the model greatly.

Agent-based models representing individuals separately can be an effective alternative to mathematical models in dealing with these issues [17]. Each individual can be represented by a particle, and interaction between particles can represent social interactions. Microsimulation modeling approaches can then be used with rules governing those interactions depending on the characteristics of the particles (such age and vaccination status). This model allows all the individuals to behave and interact according to separate characteristics and we can than observe the resulting macroscopic and microscopic impact on society.

Here we present MAM, Monte-Carlo, Agent-based Model, a novel method to model infections and disease spread. We show that MAM provides a high level of flexibility in modeling heterogeneous populations and, in turn, outperforms SIR-based models in predicting COVID-19 outbreaks in Israel. We show predictions for the number of confirmed cases and the prevalence of variants in Israel for three major outbreaks, which were dominated by different variants and different social behaviors.

## Methods

### The Monte Carlo Agent-based Model (MAM)

Here we provide a short description of MAM and a pseudo code for the algorithm. The approach is to generate a spatial model of the population by using a set of interacting classical particles, as in an agent-based model [18, 19]. We then apply standard Monte Carlo (MC) procedures of sampling the transition among subsequent states, which are sampled from a statistical distribution as in MC methods for electron transport [20-22].

For modeling the spread of the disease, each particle can be in one of four states: (i) susceptible and unprotected (unvaccinated), (ii) susceptible and protected (vaccinated), (iii) currently infected and contagious, and (iv) recovered/dead. On any given day, the probability of a susceptible particle becoming infected depends on how far it is from every infected particle around it, and whether the particle is protected. Particles move in the space and change their position every day using an MC procedure. Since the infection probability is a function of the relative distance between particles/individuals, we can use different baseline spatial characteristics, that may represent a scattered city suburb or a dense university classroom.

Moreover, the spatial features of the model allow simultaneous modeling of multiple geographic areas, each of which has unique vaccination rates and allows modeling infection for different infection circuits, such as families and close and remote communities [23].

The main difference between the SIR and MAM is that in MAM each particle can be tuned individually, as opposed to SIR, where there are limited number of subgroups that can be modeled together. In MAM, inputs such as ***R***_***t***_ (which is essentially the particle density), vaccination rates and vaccine effectiveness (VE), can be tuned for as many subgroups as needed. Therefore, this modeling approach offers high flexibility for modelling spread in populations that have different dynamics in age groups, geographical areas, etc., and in adapting the model to different VE scenarios.

### The Monte Carlo Agent-based Model (MAM) pseudocode

#### Algorithm 1

[*t*_*I*_, *H*]=MAM(*N, N*_*I*_, *N*_*Rc*_, *L*_0_, *R*_*t*_, *T*_*V*_, *T*_*span*_, *VE*_*days*_, *VE*_*level*_)

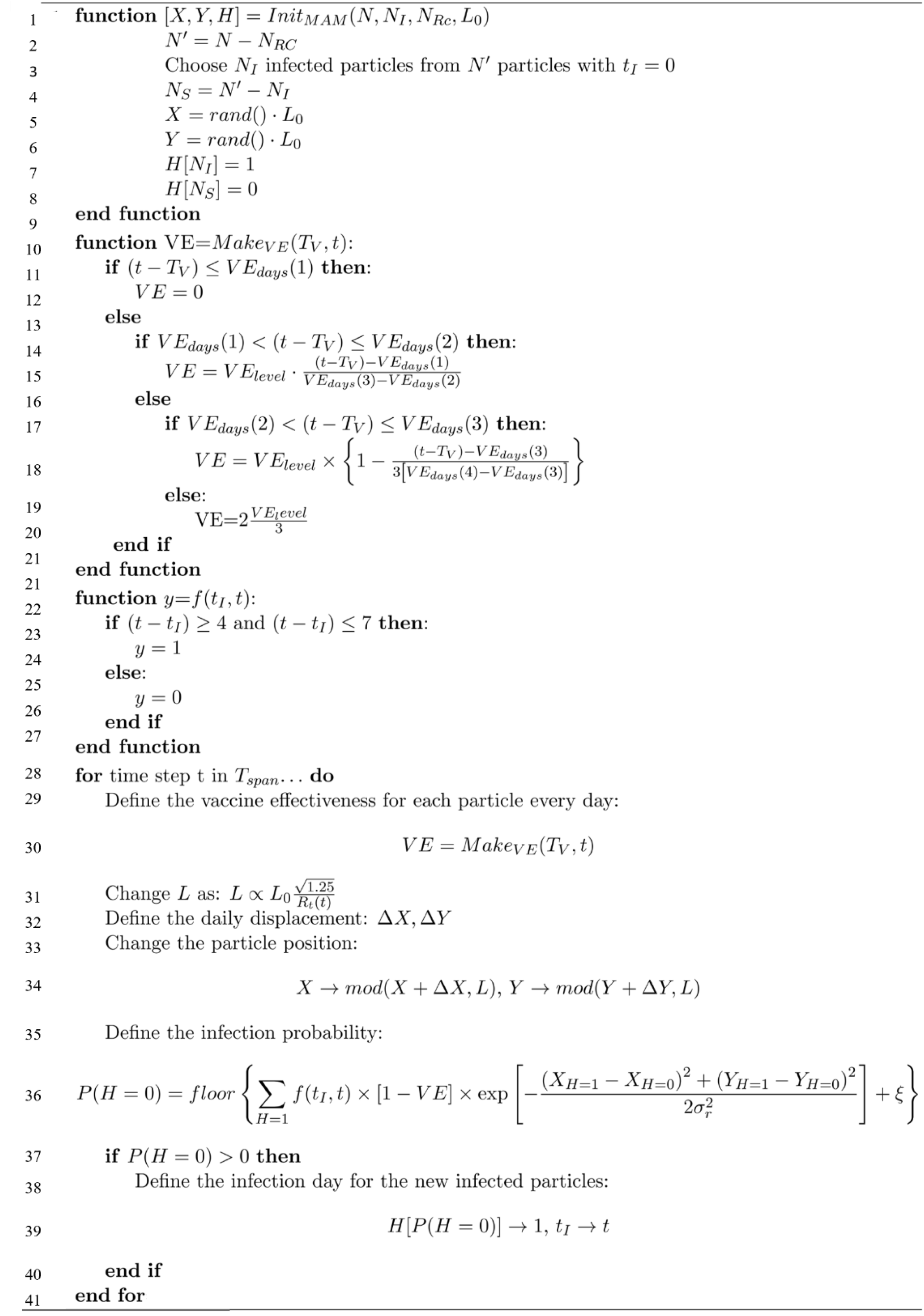

Where:

- *H* is the health status (0 for susceptible and 1 for infected) of each particle.
- *t*_*I*_ donates the particle day of infection.
- *f*(*t, t*_*I*_) is a binary function for the infection period. During the Alpha and Delta variants, the infectious period for a particle is between the fourth and seventh day after infection. During the Omicron variant, the infectious period for a particle is between the second and fifth day after infection.
- *N* is the total number of particles, *N*_*I*_ is the initial number of infected particles and *N*_*RC*_ is the initial number of recovered particles. Here the simulation is set with 1.1 ⋅ 10^4^ particles, in which the serial number of the particle denotes its age, i.e., the top 17% numbers are the 60+ population, and the bottom 20% numbers are the 0-11 population. For simulating the population in Israel (9.2 ⋅ 10^6^ people), the simulation runs ∼ 800 times.
- *L*_0_ is the initial length of the simulation area, *A*, such that ***R***_***t***_ is represented by the particles’ density. For the case of *N* = 1.1 ⋅ 10^4^ particles we find that for ***R***_***t***_ ≈ 1.25, *L* = *L*_0_ = 124 meter and 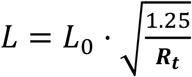.
- *T*_*V*_ indicates the day of immunization for each particle (*T*_*V*_ = ∞ for unvaccinated particles).
- We us here actual daily immunization rate by age groups taken from https://data.gov.il/dataset/covid-19.
- *T*_*span*_ is the simulation duration time
- *VE*_*days*_ are the days after the first dose when vaccine effectiveness changes with *VE*_*level*_ being the maximal vaccine effectiveness, i.e., for this work *VE*_*days*_(1) = 7 days, *VE*_*days*_(2) = 28 days, *VE*_*days*_ (3) = 150 days and *VE*_*days*_(4) = 180 days with *VE*_*level*_ = 90% (see **Figure S2 A**).
- *X* and *Y* are the daily x and y positions for each particle where Δ*x* and Δ*y* distribute. normally with *μ* = 0 and 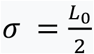. For each direction, we apply a periodic boundary condition such that: 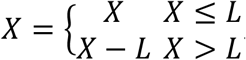.

### Modelling COVID-19 spreads in Israel for three outbreaks

To model the spread, for both SIR and MAM models, there are several parameters that must be tuned. The main metric required for modelling the spread of COVID-19 is ***R***_***t***_, the theoretical reproduction rate of the disease, which is an estimation of the average amount of encounters between a carrier and healthy individuals that would have resulted in an infection if the vaccines had not been available. ***R***_***t***_ is a function of the level of infectiousness of the variant, ***R***_**0**_ (the virus basic reproduction rate) and NPIs used to control the spread of the disease, notably mandatory isolation for individuals with confirmed infections and those exposed to a confirmed case, and face mask-covering indoors. It is thus a dynamic metric, that changed throughout 2021 with relaxing and stringing restrictions, and the introduction of new variants (**Figure S1A**). The Alpha variant, which was dominant in Israel in the beginning of 2021, had an ***R***_**0**_ of approximately 4, while the Delta variant, which became prevalent in Israel at the beginning of May 2021, had an ***R***_**0**_ of 5-8 (**Figure S1B**) [24]. It should be noted that testing-availability and testing-policy vary not only by country but also by time (**Figure S1C**), and thus, confirmed cases is a problematic metric for comparing disease spread across countries and across time. In MAM, ***R***_***t***_ is represented by the particle density or the size of the area for the particles (*A*). In this work, we estimate ***R***_***t***_ in Israel from January 2021 to February 2021 (the first outbreak) to be 1.2, similarly to a previous study we performed [25]. For the case of the delta and omicron outbreaks, as a result of the removal of most social restrictions in Israel, we estimate ***R***_***t***_ for the delta and omicron outbreaks to be 2.2-2.5 (see Table S1 and S2).

For modelling with SIR, we also need to tune ***R***_***e***_. ***R***_***e***_ reflects not only ***R***_**0**_ and the NPIs used to control disease spread but also the population vaccination rate and the vaccine effectiveness (VE) in protecting against infection. Therefore, to tune ***R***_***e***_ we need to have an estimation of VE, which might be non-trivial as it is also a dynamic measure that changes over time. Here, VE was set to 0% on days 0-7 after the first dose of vaccination and increases linearly up to 90% on the next 21 days, which is also one week after the second dose (**Figure S2A**) [26, 27]. We next assume reduction to 60% 180 days later, and following the booster dose (third dose), we assume increase back to 90% seven days after the vaccination. Since ***R***_***e***_ is a function of vaccination rates that change over time, for each age group, we need to define a time-dependent ***R***_***e***_ which is a function of both protection level as a function of time and of the vaccination rate of the relevant age group. In MAM, no need to estimate ***R***_***e***_, but VE is provided as input.

Vaccination rates differ among different populations and age groups (**Figure S2B**). In Israel, by the end of 2020, in parallel with a major COVID-19 outbreak, a massive vaccination campaign was initiated. Elderly individuals were offered to vaccinate first, and every couple of weeks the age limit was reduced. By early January 2021, over 60% of those 60 years old and over were vaccinated with at least one dose. It was later offered to younger individuals to vaccinate, but this was a much slower process. This exemplifies, that ***R***_***e***_ cannot be tuned correctly to the whole population but is a measurement that is specific to subgroups of the population.

In this work, for modeling the spread, we first used a “naïve” SIR model, which assumes homogeneous protection for all the population without age-based divisions. Next, we used a multi-age SIR [30, 29] and MAM models such that we divided the population into three age groups: Group A represents the 0-11 years old population, which is 20% of the population who was not vaccinated until the end of 2021; Group B represents the 12-59 years old population, which is 63% of the population; and Group C which represents the 60 years old and over population, which is 17% of the population (**Figure S2A**).

To exemplify how this affects modeling, assume a theoretical case that the entire age group is vaccinated on Feb 1st. We would expect that after 28 days, the average protection against infection of that age group would be 90% (**Figure S2B**), and therefore, their chance of infection is 10%. As a result, effective reproduction rates should vary for each group based on vaccination rates (**Figure S2C-D**), despite the assumption that populations mix homogeneously, and everyone is susceptible to infection without vaccinations (**Figure S2C-D**). Therefore, we have a system of 9 coupled differential equations (3 health status for each of three age groups) which we need to solve simultaneously (**Supplementary Data 1**) [30].

In the MAM model, all the particles have a serial number, and thus the population is divided into subgroups based on the particle rank using similar proportions: the 17% highest ranked particles represent the 60+ age group, the next 63% represent group B, and the last 20% particles represent group A based on the age distribution in Israel [31].

Both models were used to predict the daily confirmed cases of three major outbreaks: December 1, 2020, to February 20, 2021 (third outbreak in Israel, Alpha); July 15, 2021, to October 31 (Delta) and Jan 1st, 2022, to March 1st, 2022. In both models, the population was divided into three age groups, and for all outbreaks, the vaccination rates inputs were obtained from the Ministry of Health until January 8 only (https://data.gov.il/dataset/covid-19). In all simulations, starting January 8 and on, we use different assumptions on vaccination rates and not the actual vaccination rates.

### Statistical analysis

To provide statistical measurement for the accuracy of the modeling predictions compared to the actual real-world data, we used the mean absolute percentage error (MAPE) [32]. This relative error measure uses absolute values to prevent positive and negative errors from canceling each other out and uses relative errors to compare forecast accuracy between time-series models. For each model, we examine the accuracy of the prediction in three age groups: the total population, the youngest and unvaccinated population, and the elderly population.

### Error estimation

To provide sample-distribution error in MAM we use the relationship between ***R***_***t***_ and *A*, the simulation area. This relationship is as follows: 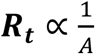, where ***R***_***t***_ is an input to the simulation (see P2). We add 10% uncertainty error for MAM. However, MAPE values for the MAM model were calculated using its mean value.

### Data

All the data that was used in this work was taken from public data sources. Daily confirmed cases in Israel was taken from https://datadashboard.health.gov.il/COVID-19/general and vaccination rates were taken from https://data.gov.il/dataset/covid-19, all stratified by age groups and available from the Ministry of Health from December 2020 to February 2022. In Addition, for the Omicron outbreak, we used data from https://covariants.org/ to examine the prediction for variants distribution over time.

## Results

### Outbreak 1: First vaccination campaign in Israel (Alpha variant)

We present the predictions of SIR and MAM modelling of the COVID-19 outbreak in Israel that started in mid-December 2020, parallel to the vaccination campaign. The Alpha variant dominated this outbreak. Data on confirmed cases and vaccination rates were collected until January 8, 2021 (dashed horizontal line in **Figure 1**), and predictions were made from this day forward. Vaccination rates from this date forward were estimated using an exponential function (**Figure 1A**). All the details for calculating this outbreak for both SIR and MAM models are given in Table S1.

**Figure 1.**
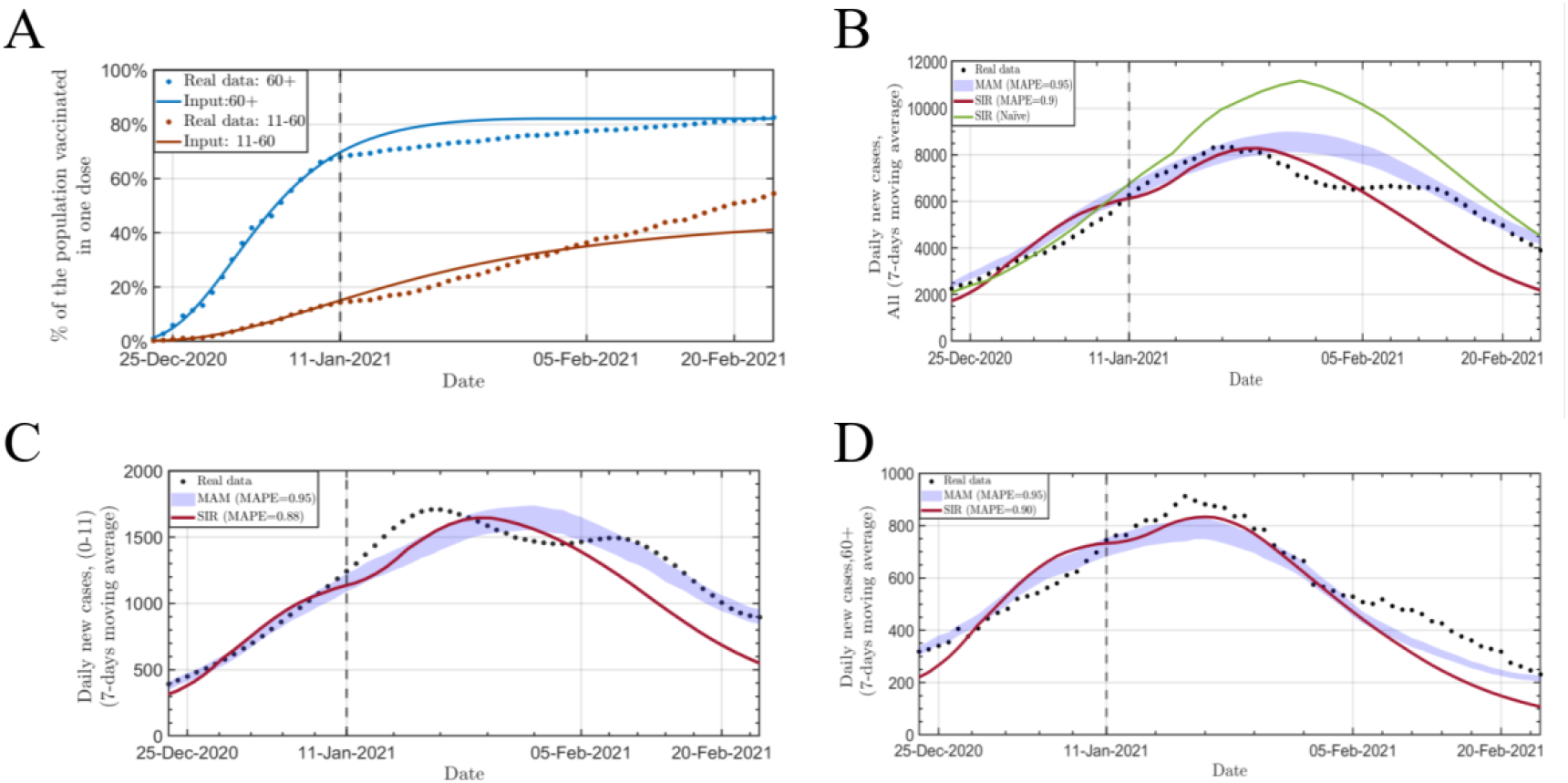
Modelling the outbreak spread in Israel during the first vaccination campaign. **A**. Vaccination rates for two age groups based on real data up to January 8th, 2021, and predicted rates from this day forward. Actual vaccination rates are in dots. **B-D**. Predictions for the number of confirmed cases in Israel from Dec 1st, 2020, until Feb 20, 2021 (7-days moving average) for the whole population (**B**), ages 0-11, which are all unvaccinated currently (**C**); and for 60+ years old (**D**). For all panels: real data in dots, SIR predictions in solid red line and MAM predictions in blue band. Solid black line on January 8th, 2021, shows the index date from which predictions begin. Green line in **B** is a prediction made with SIR without breaking down age groups. MAPE statistics are presented in the legend.

First, we apply a naïve SIR model, assuming average protection of the total population (**Figure 1B**). Interestingly, a naive SIR approach fails and overestimates cases since the average protection of the total population is lower than the average protection for each vaccinated individual, as it considers the unvaccinated. This causes ***R***_***e***_ to be overestimated, and in turn overestimate the number of expected confirmed cases.

We, therefore, applied multi-age SIR and MAM models to predict the number of confirmed cases from January 9th to mid-February 2021 in three age groups (**Figure 1B-D**). Both SIR and MAM models were able to accurately predict the peak, the date when the number of confirmed cases peaked and started to decline (***R***_***e***_ < 1). However, MAM consistently outperformed SIR in predicting the decline in all three predictions. Across all three analyses, MAM outperformed SIR in terms of accuracy (MAPE = 84%-86% for SIR versus 93% for MAM).

### Outbreak 2: Waning immunity and booster dose (Delta variant)

This outbreak, which started in June 2021, was characterized by three factors that overlapped: relaxation of NPIs, the introduction of the more infectious Delta variant, and waning immunity half a year after the first vaccination campaign. The combination of all the above resulted in a higher *R*_*t*_ than that of the first outbreak [33]. On July 31, 2021, Israel initiated another vaccination campaign for a booster dose, which similarly took place to the first campaign: the elderly population first, and later opened it for the rest of the population.

Here, we modeled the outbreak according to information gathered until August 1st, 2021, as the vaccination campaign has just started. Since vaccination dynamics were unknown then, we assumed similar dynamics, by age groups, as in the first vaccination campaign (**Figure 2A**). We first applied the naive SIR model to the whole population, assuming only the average protection for the whole population (**Figure 2B**). As a result of the assumption that the unvaccinated population has a higher ***R***_***e***_ than that of the first outbreak, the number of daily confirmed cases was significantly overestimated. This significant deviation results from the exponential nature of pandemic spread, a 10% increase in ***R***_***e***_ can result in a 50% increase in the number of total cases. It is clear that a naive model cannot be applied to accurately predict the spread in scenarios where the infection potential for the unvaccinated is significantly higher than for the vaccinated. (All the details for calculating this outbreak for both SIR and MAM models are given in Table S2).

**Figure 2.**
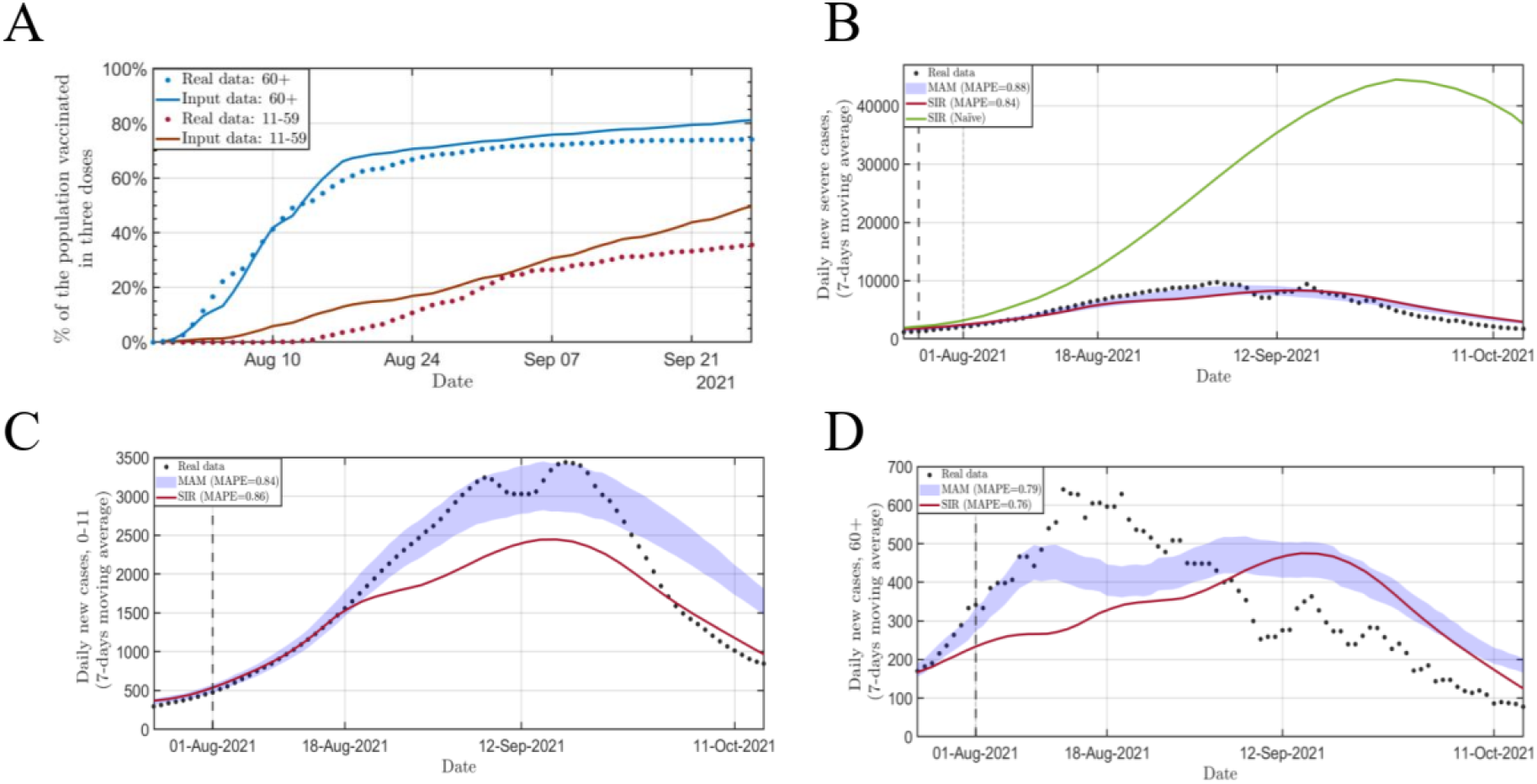
Modelling the outbreak spread in Israel during the booster vaccination campaign. **A**. Vaccination rates of the booster dose for two age groups based on real data and predictions from August 1st, 2021. Actual vaccination rates are in dots. **B-D**. Predictions for the number of confirmed cases in Israel from July 21st, 2021, until Oct 30, 2021 (7-days moving average) for the whole population (**B**), ages 0-11, which are all unvaccinated at this time (**C**); and for 60+ years old (**D**). For all panels: real data in dots, SIR predictions in solid red line and MAM predictions in blue band. Solid black line on August 1st, 2021, shows the index date from which predictions begin. Green line in **B** is a prediction made with SIR without breaking down age groups. MAPE statistics are presented in the legend.

Both multi-age SIR and MAM models were able to predict the dynamics of this outbreak well, yet MAM predicted the peak better and had an overall accuracy of 88% compared to 84% with SIR (**Figure 2B**). At the young age group (ages 0-11), MAM accurately predicted the peak, but compared to the real data, the decrease of this outbreak was much sharper than predicted (**Figure 2C**). A possible explanation is that the peak of the outbreak intersected with the Jewish high-holidays, which is a time of school closing, and later, a program of mandatory rapid testing before return to school, and both together made the outbreak subside quickly in young children. Both models did not perform well in the age group of 60+ years old, but both had relatively good estimation for the end of this outbreak. Similar to all other comparisons, MAM was more accurate than SIR here as well (**Figure 2D**).

### Outbreak 3: Competing variants

This outbreak, which started in December 2021, was characterized by the highly infectious Omicron variant. This variant rapidly became dominant, with individuals arriving from abroad with Omicron, while Delta was also on the rise. It is important to note that Delta and Omicron differ significantly in their ***R***_**0**_: here we assume that Omicron ***R***_**0**_ is double of Delta’s ***R***_**0**_ and in addition, we assume a shorter incubation time for Omicron (**Table S3**) [34, 35]. A similar scenario happened later on in February, where Omicron BA.1 was replaced by the BA.2 variant, again because of higher ***R***_**0**_ (we assume here 50% higher ***R***_**0**_ for BA.2 compared to BA.1) [36].

It is unclear how to model such situations with multiple ***R***_**0**_ and different incubation times with SIR; therefore, the variants analyses are modelled with MAM solely.

Using data on number of infected individuals that arrived in Israel in the first half of December (assuming all are Omicron) and number of confirmed cases in Israel in that time frame (assuming all are Delta), we attempted to predict the proportion of the variants in the next 30 days. By using MAM modelling, where each particle represents an individual, such a task is feasible, and MAM was able to accurately predict the dynamics of variants transition (**Figure 3A**). We also repeated this modelling for the BA.1-BA.2 transition, using data of influx from January 19, 2022, to the mid of February, and predicted variants distributions in February-mid till April 2022 (**Figure 3B**).

**Figure 3.**
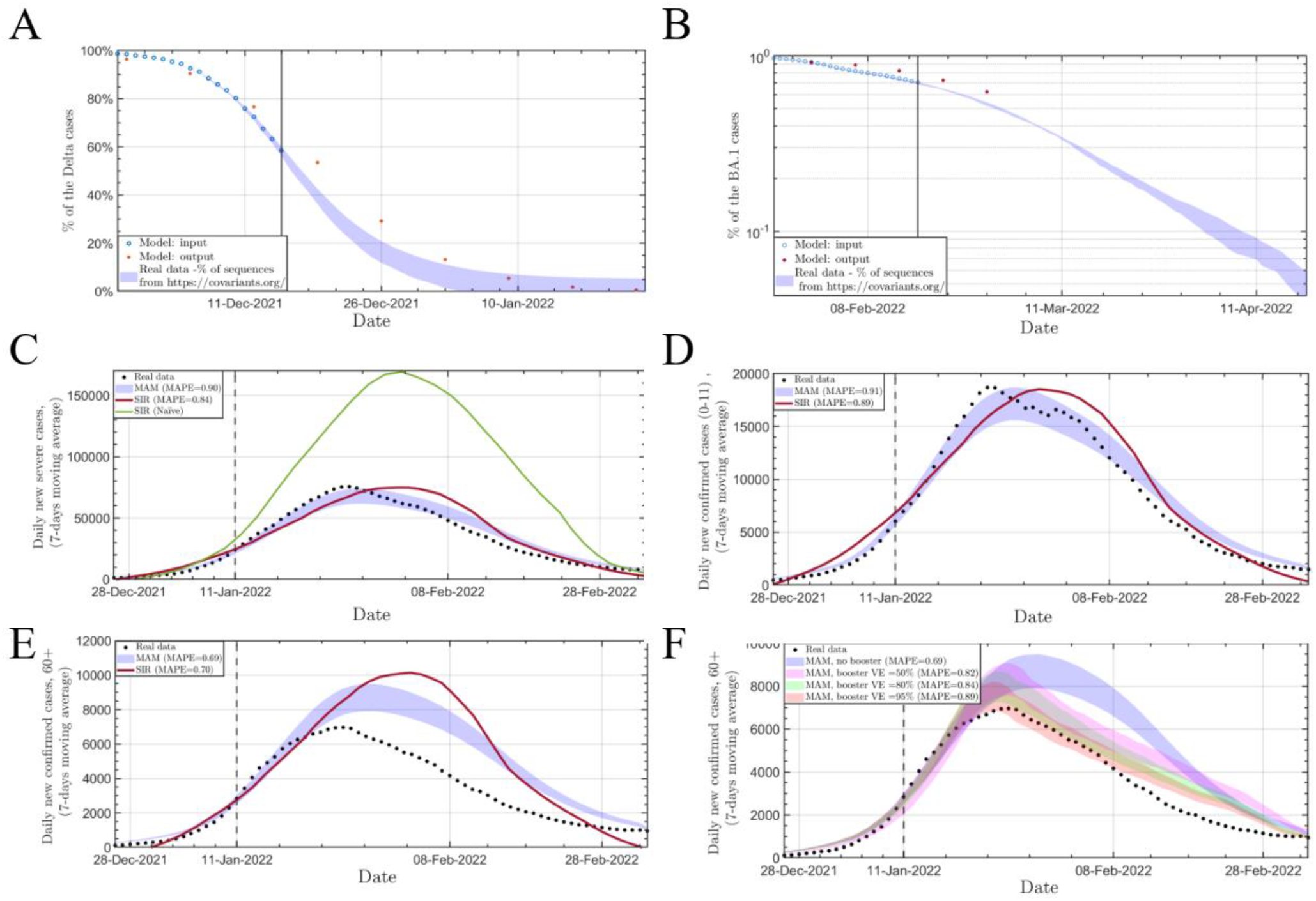
Modelling the outbreak spread in Israel during the Omicron outbreak. **A**. Percentages of Delta variant from December 1st, 2021, and prediction with MAM from December 15th, 2021, as it was replaced by Omicron. Red dots are actual data. **B**. Percentages of Omicron BA.1 variant from January 19th, 2022, and prediction with MAM from February 15th, 2022, as it was replaced by the BA.2 variant. Red dots are actual data. **C-F**. Predictions for the number of confirmed cases in Israel from January 1st, 2022, until February 28, 2022 (7-days moving average) for the whole population (**C**), ages 0-11, which are all unvaccinated at this time (**D**); for 60+ years old (**E**); for 60+ years old with knowledge on the protective effect of dose 4 for three different vaccine effectiveness against infection **(F)**. For all panels: real data in dots, SIR predictions in solid red line and MAM predictions in blue band. Solid black line on January 11th, 2022, shows the index date from which predictions begin. Green line in **C** is a prediction made with SIR without breaking down age groups. MAPE statistics are presented in the legend. For **F**, the magenta, green and red bands are the MAM model with 50%, 80% and 90% vaccine effectiveness, respectively, against infection for the fourth dose.

We next used data collected up to January 11, 2022, to model the outbreak from then forward, and predict daily confirmed cases under the assumption that from that day and on, Omicron is the dominant variant, which enable the use of the multi-age SIR model. Based on the data gathered up to the index date, we assumed here ***R***_***t***_ = 2.5. In MAM, since there are competing variants, we also assumed that Omicron is twice is infectious as Delta.

Similar to the previous outbreaks, we first employ the naive SIR model (**Figure 3C**). The naive model again overestimated the actual data, but not extreme overestimation as in in outbreak 2. This is because in this outbreak the vaccinations provided only little protection. We next applied MAM and multi-age SIR to the three age groups. Both models successfully predicted the number of cases in the whole population and the young populations, with MAM predicting better the peak and with better MAPE (**Figure 3C-D**). However, both models did not predict well the peak and the rapid decrease in cases in the 60+ year old population (**Figure 3E**).

On January 2, 2022, Israel started another vaccination campaign, this time for a fourth dose, and this time only for individuals over 60 years old. By January 10, 2022, 15% of the 60+ year-old population received the fourth does, and two weeks later, this figure went up to about a quarter. Based on the deviation between our predictions and the number of daily confirmed cases, which begun after January 18^th^, we hypothesized that this vaccination campaign was the primary reason for the early drop in cases. It should be noted that there is yet a debate of how effective this campaign was and whether it had an effect and the outbreak. Also, other possible explanation for this deviation between the prediction and the actual data is that the older population have taken extra precautions during a major outbreak.

There have been several empirical studies to determine the effectiveness of the fourth vaccine dose against infection [37, 38], which found that for there is the effectiveness of the fourth dose against infection is around 50%, which waned in later weeks. Here, we attempted to predict VE just from vaccination rates and aggregated data of confirmed cases. We performed MAM predictions for the 60+ population using three different VE levels and found that the best fit to the actual data was with *VE* = 95%, which is much higher that the estimations received from rigorous studies using individual-level data (**Figure 3F**). Nevertheless, a model that assumes 50% effectiveness of the fourth dose shows a significant decline in confirmed cases compared to a non-booster scenario (**Figure 3F**). Hence, the model results, actual data, and effectiveness tests of the fourth dose suggest that the older population took additional precautions to prevent infection. This, together with the effectiveness of the fourth dose, resulted in a lower incidence of morbidity for this population.

Finally, an important question is how changes in ***R***_***t***_, possibly by enforcement of more stringent NPIs, may affect the dynamics of the outbreaks. We used MAM to compare how a reduction of 20% in ***R***_***t***_ will affect the number of daily confirmed cases and, possibly more importantly, the number of total severe cases in hospitals in parallel. This was the primary metric used by the government to decide whether enforcing stringent NPIs is required, as the main goal was to prevent hospital overflow. We compared three scenarios: A) no change in ***R***_***t***_ ; B) Temporary reduction in ***R***_***t***_ for two weeks starting January 15, which may reflect two-weeks school closure and C) Reduction of ***R***_***t***_ by 20% from January 15 (**Figure 4A-B**). For scenario C, the analyses predicted 40% fewer total infections and severe cases from January 15 until February 28 in comparison to scenario A (**Figure 4A-B**). Importantly, in both scenarios B and C, our prediction suggested that the peak of active severe cases would not have crossed 900 severe cases in parallel (**Figure 4B**), which was the limit in Israel before a significant decrease in the level of care [39].

**Figure 4.**
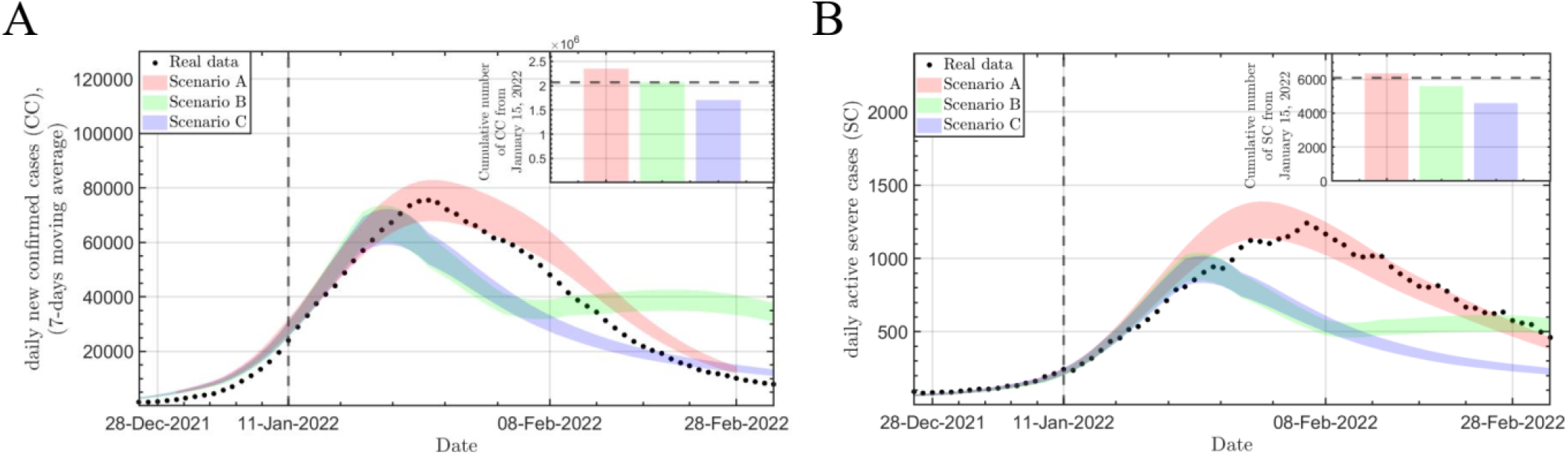
Comparing effects of alternative scenarios of enforcing restrictions. **A-B**. Comparison of predictions for three scenarios for the daily number of confirmed cases (**A**) and parallel severe cases (**B**) in Israel from January 1^st^, 2022, until February 28, 2022 (7-days moving average) for the whole population. Scenario A is no change in Rt, scenario B is temporary 20% reduction in Rt for two week and scenario C is 20% reduction in **R**_**t**_ s. Solid black line on January 11^th^,, 2022, shows the index date from which predictions begin. For both panels, the inner plot is the cumulative number of cases from Jan 15^th^ t until Feb 28 for the three scenarios. The dashed line is the actual data taken from https://data.gov.il/dataset/covid-19.

Another insight from these analyses is the divergence that we observe between the actual data and predictions from around January 24. We only observe sharp reduction compared to the prediction in confirmed cases, but not in severe cases. During that time, testing policy in Israel changed, and there was arguably undercount of number of actual cases. However, since the number of severe cases is not affected by changes in testing policies, and therefore the model was more accurate in that prediction.

## Discussion

We learned over the past two and a half years that the reality of SARS-CoV-2 is rapidly changing, with the introduction of numerous variants, each with its own unique characteristics, vaccination campaigns, waning immunity, and ever-changing decisions regarding NPIs and testing requirements. For this reason, modeling disease dynamics requires a dynamic model that can take all these factors into account. We presented here a novel particle-based approach to model disease outbreaks, which we named MAM, that provides high flexibility in modelling such complex situations. We showed its superiority over the widely used multi-age SIR model in predicting the dynamics of outbreaks in Israel. It should be noted that we used similar assumptions for SIR and MAM in all the analyses. We also showed here that MAM is highly flexible, allowing to easily define different vaccination rates in subgroups of the population, changes in NPIs, and competing variants.

We found that a naive SIR model significantly overestimates outbreaks, especially when vaccines are in effect. This could be by the Simpson paradox, as vaccines are effective on the individual level, but when merging all age groups together, which have different vaccination rates, the average protective effect seems low. Another reason for this failure is that when vaccines provide high level of protection, unvaccinated individuals benefit from the indirect protection [40, 41]. In Israel, since vaccination campaigns were conducted by age groups, there was cumulative protection in groups that were vaccinated early in addition to the protection from the vaccine itself. This, an assumption of uniform protection across the population is not sufficient to correctly model the spread of the disease. For these reasons we applied a multi-age approach using both SIR and MAM. In this approach each age group has its own characteristics based on vaccine effectiveness and vaccination rate, and assuming homogenous mixing across the three age groups. Both approaches provided relatively good predictions in the three outbreaks we modeled from Israel; however, MAM was consistently more accurate. It is important to note that here we used only three age groups and not more, as adding additional age groups to SIR is complex and prawn to errors. In MAM, this is straightforward.

Another complication we introduced here is modelling outbreaks when there are competing variants. We showed here that such situations can be modeled efficiently from the very early stage of the outbreak with MAM, since we can provide each particle different adhesion characteristics, such as adhesion potential and incubation time.

Finally, dynamics of infection can vary across different groups and geographies. The MAM approach has a built-in spatial structure since the particles move in space and change their position with average daily displacement. Thus, adding spatial characteristic, such as households, neighborhoods, cities etc. can easily be applied to this model, and provide additional precision to modelling the disease spread.

In summary, we presented here a novel approach for modelling the spread of the pandemic and exemplified its accuracy and flexibility across different scenarios. We hope that this tool will be utilized in the future to make informed decision-making on how to deal with disease outbreaks

## Data Availability

The code is available in the text and the data is taken from https://datadashboard.health.gov.il/COVID-19/general and https://data.gov.il/dataset/covid-19

https://datadashboard.health.gov.il/COVID-19/general

## Supplementary materials

### Supplementary figures

**Figure S1.**
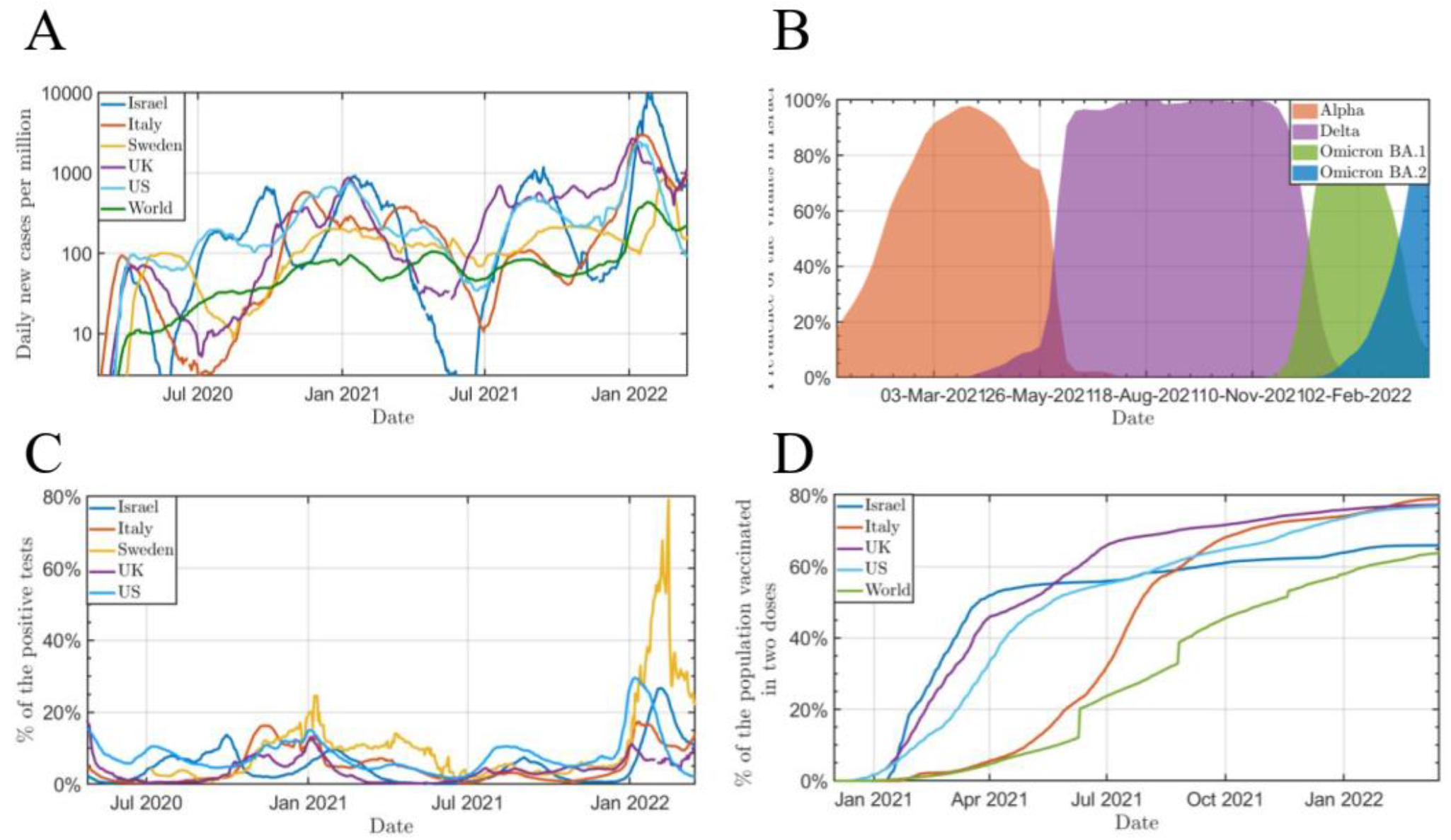
Time-dependent features of COVID-19 spread and detection. Panel A: The normalized number of confirmed cases as a function of time in various countries (data was taken from https://ourworldindata.org/covid-cases). Panels B: Variants of COVID-19 in Israel over time (data was taken from https://covariants.org/). Panel C: Percentage of the daily tested population (data was taken from https://ourworldindata.org/covid-cases). Panel D: The percentage of people who have received at least two doses of vaccination (data was taken from https://ourworldindata.org/covid-cases).

**Figure S2.**
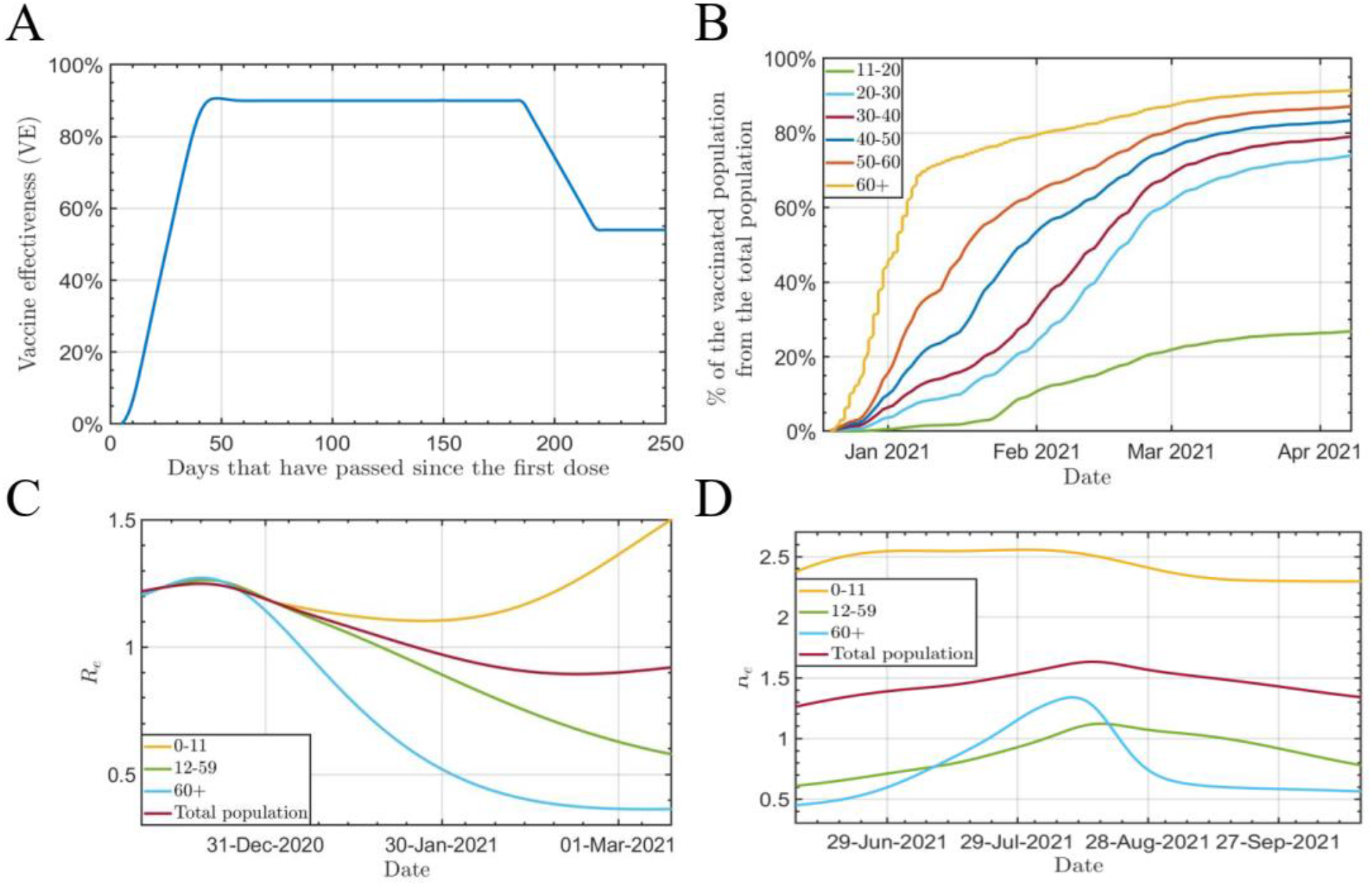
Data needed for modeling COVID-19 in Israel. Panel A: Vaccine effectiveness as function of time [27, 28]. Panels B: The rate of vaccination in Israel for different age groups (data was taken from https://data.gov.il/dataset/covid-19). Panel C and D: The R_e_ was used to model the spread of COVID-19 in Israel using naive and multi-age SIR models.

### Supplementary tables

**Table S1.**
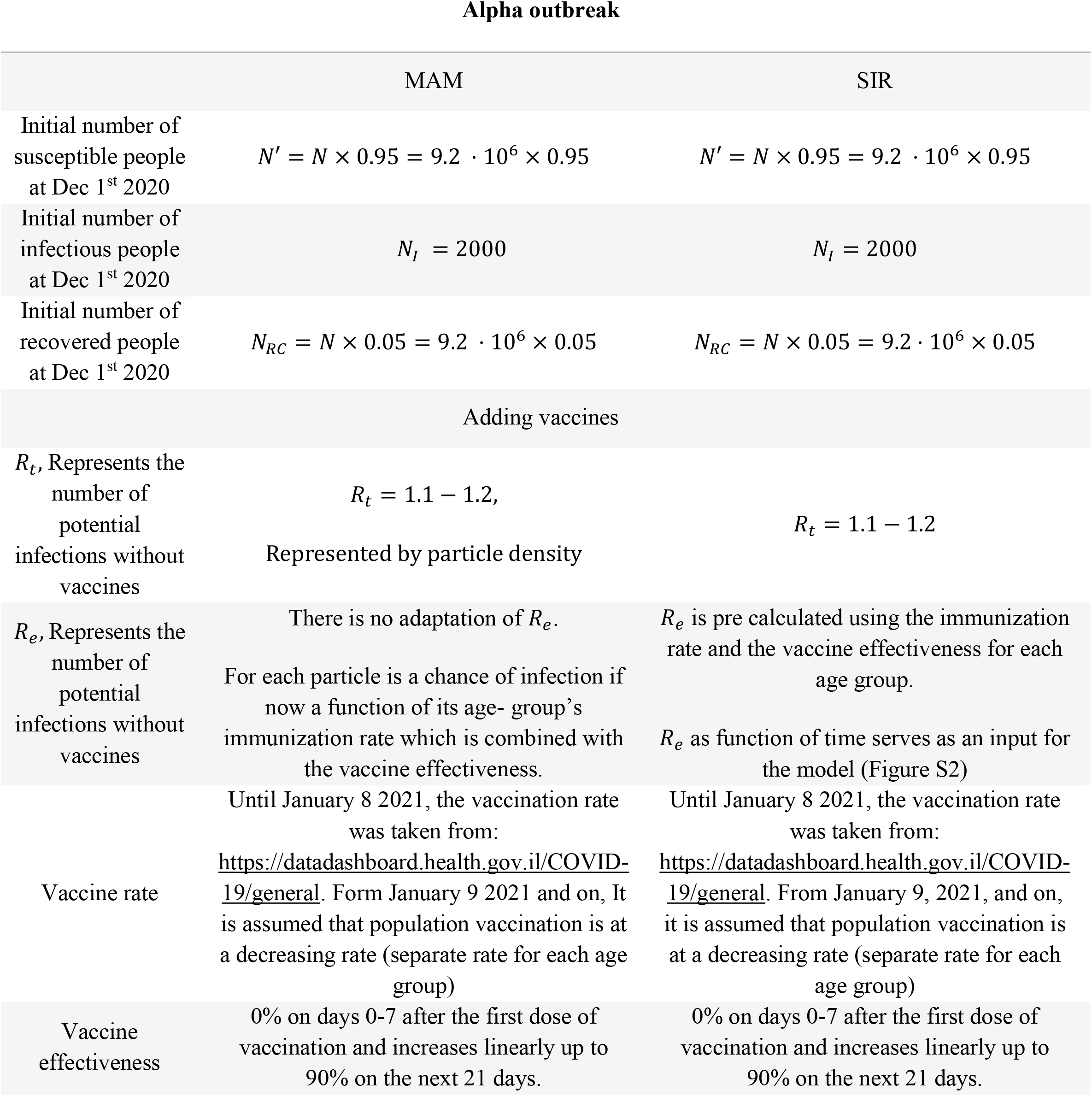
Parameters needed for modeling the Alpha outbreak in Israel for both sir and MAM models.

**Table S2.**
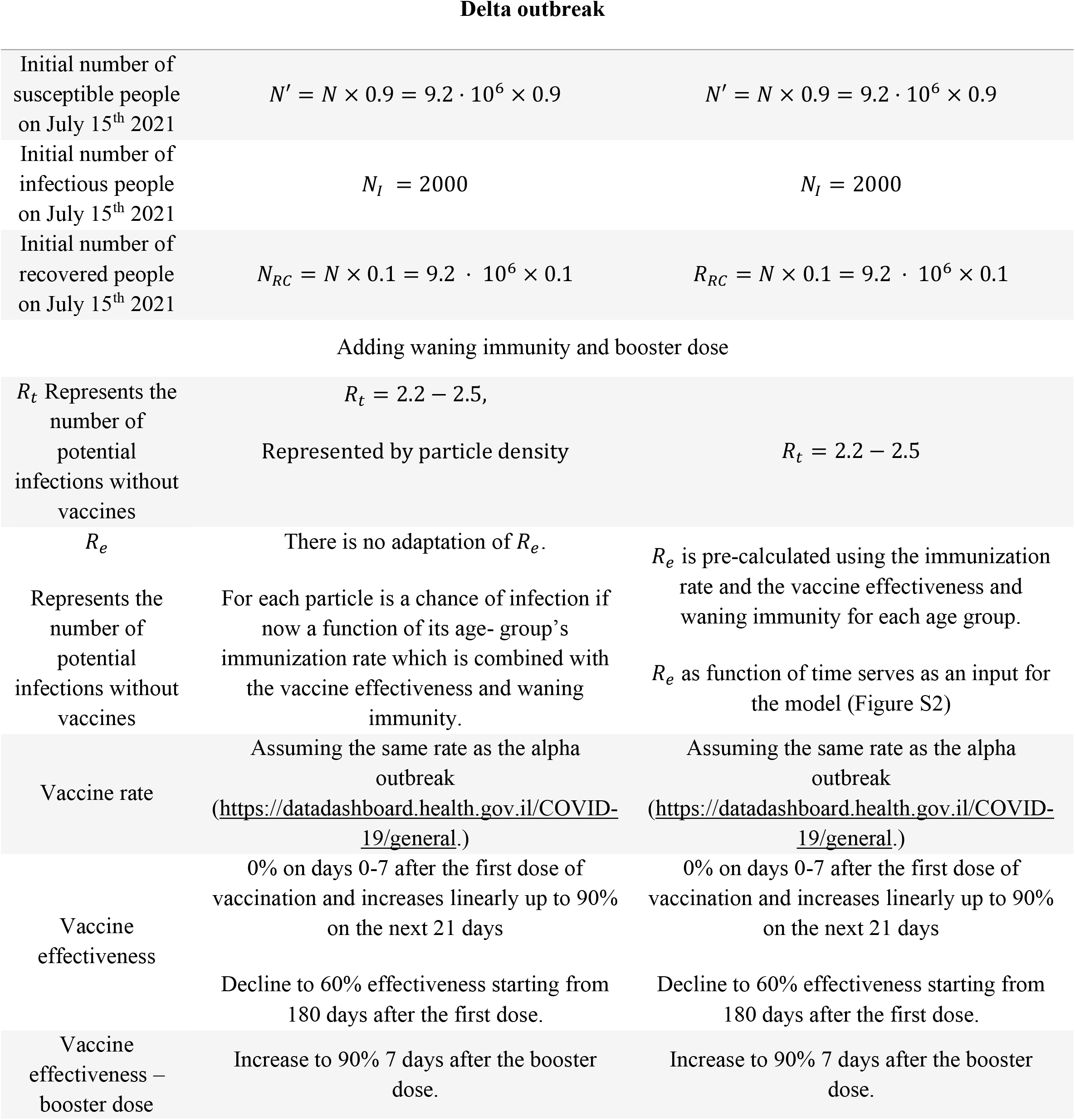
Parameters needed for modeling the Alpha outbreak in Israel for both sir and MAM models.

**Table S3.**
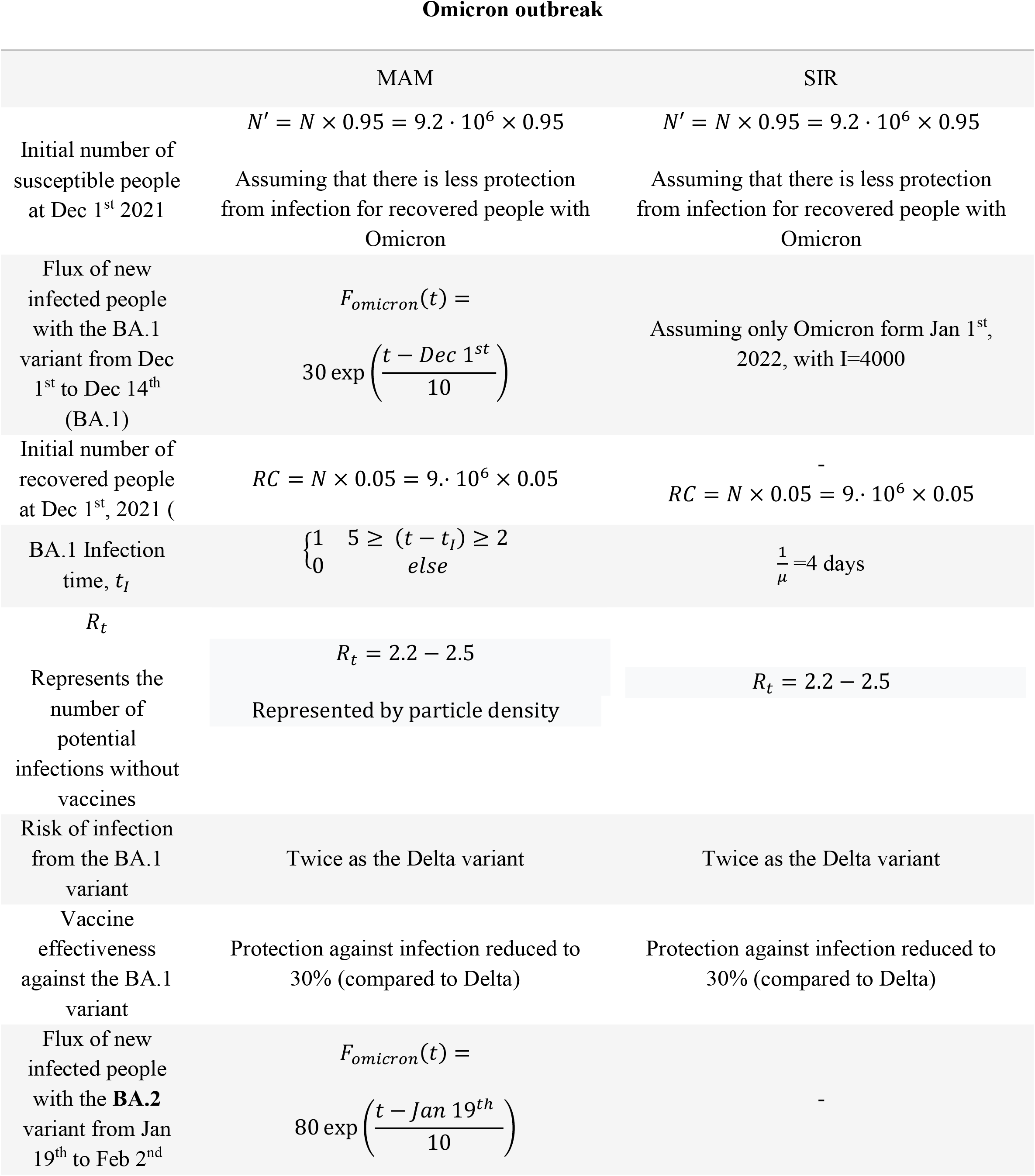
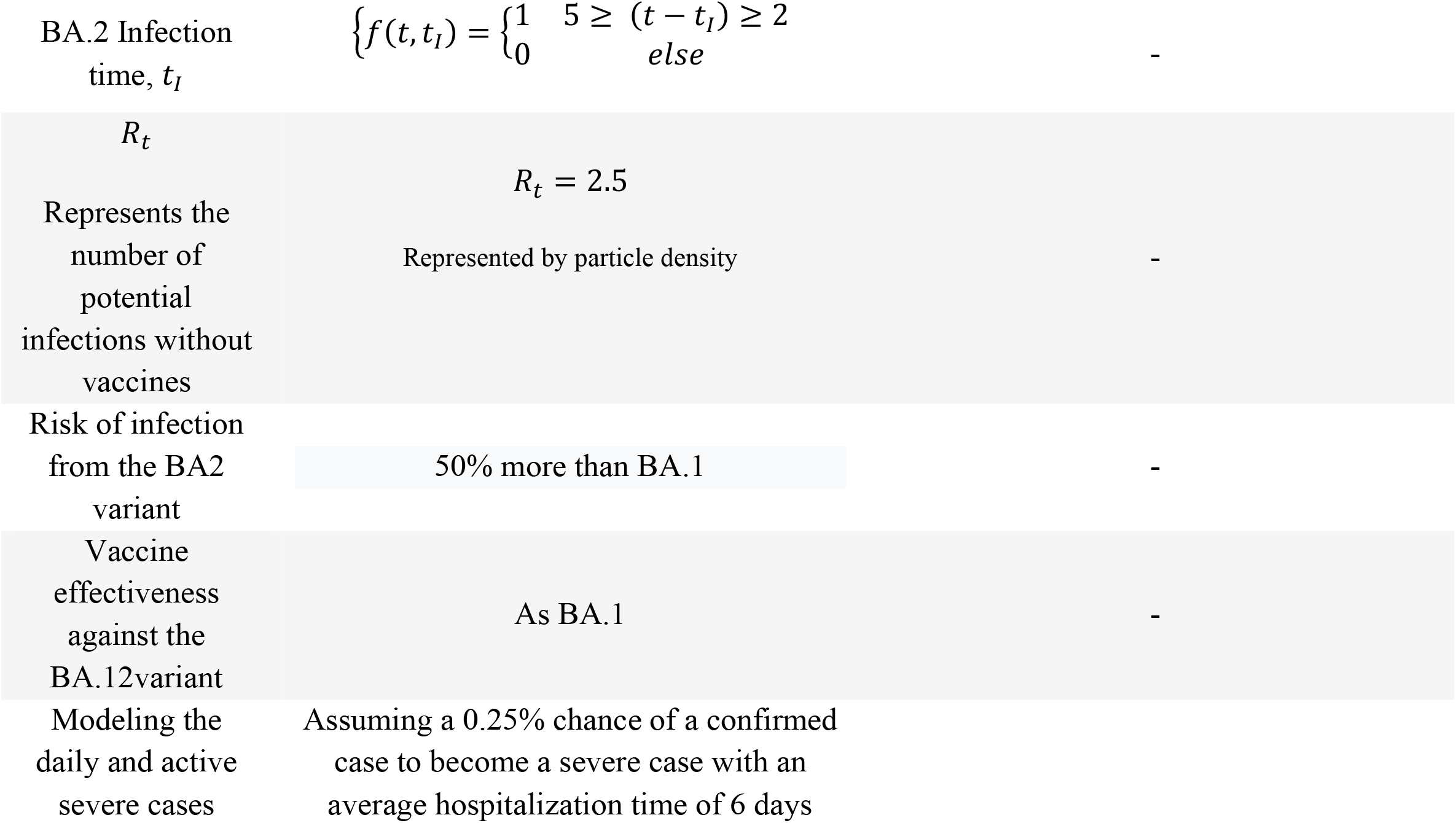
Parameters needed for modeling the Alpha outbreak in Israel for both sir and MAM models.

### Supplementary data - pseudocode for multiage SIR model

**Supplementary data 1:**
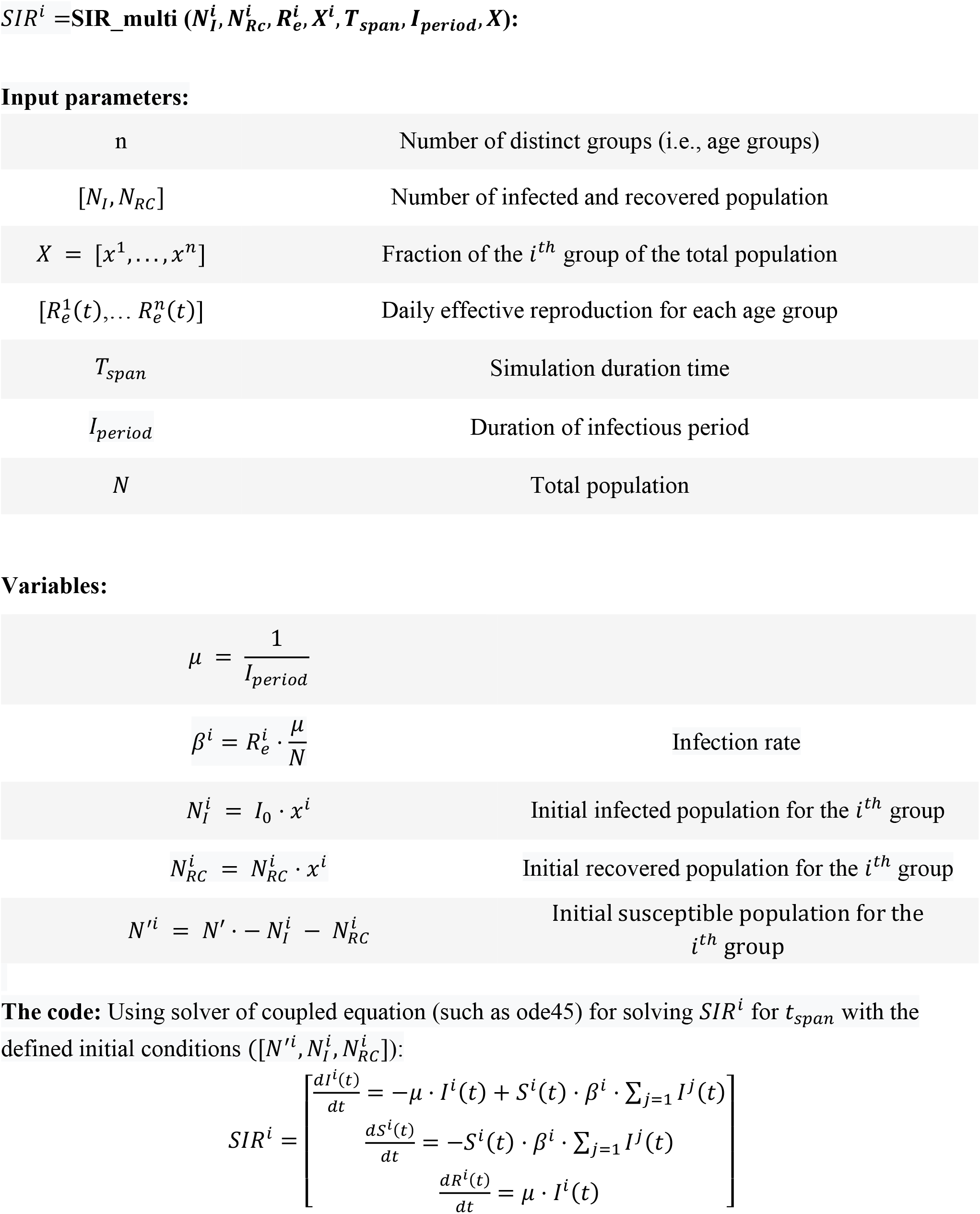
Pseudocode for multiage SIR model

Input parameters:
|  |  |
| --- | --- |
| $n$ | Number of distinct groups (i.e., age groups) |
| $[N_I, N_{RC}]$ | Number of infected and recovered population |
| $X = [x^1, \dots, x^n]$ | Fraction of the $i^{th}$ group of the total population |
| $[R_e^1(t), \dots, R_e^n(t)]$ | Daily effective reproduction for each age group |
| $T_{span}$ | Simulation duration time |
| $I_{period}$ | Duration of infectious period |
| $N$ | Total population |
| Variables: |  |
| $\mu = \frac{1}{I_{period}}$ | |
| $\beta^i = R_e^i \cdot \frac{\mu}{N}$ | Infection rate |
| $N_I^i = I_0 \cdot x^i$ | Initial infected population for the $i^{th}$ group |
| $N_{RC}^i = N_{RC}^i \cdot x^i$ | Initial recovered population for the $i^{th}$ group |
| $N'^i = N' - N_I^i - N_{RC}^i$ | Initial susceptible population for the $i^{th}$ group |

**The code:** Using solver of coupled equation (such as ode45) for solving $SIR^i$ for $t_{span}$ with the defined initial conditions ( $[N'^i, N_I^i, N_{RC}^i]$ ):

## Notes

### Competing Interest Statement

The authors have declared no competing interest.

### Funding Statement

This study did not receive any funding

